# Enhancing Access to and Utilization of Maternal and Child Health Services through Implementation of a District-led Integrated Mobile Health Service Delivery Model in Pastoral Ethiopia: A Multi-Method Implementation Research

**DOI:** 10.64898/2026.11.18.26368458

**Authors:** Agumasie Semahegn, Mebrie Belete, Gizachew Tadele Tiruneh, Yibeltal Siraneh, Shegaw Mulu, Mikiyas Teferi, Addis Girma, Yenegeta Walelign, Muluken Dessalegn Muluneh, Aklog Getnet, Mariamawit Asfaw, Mezgebu Kebede, Lidiya Tefera, Kidest Lulu Hagos, Temesgen Ayehu, Yibeltal Kiflie Alemayehu, Desalew Emaway, Lisanu Taddesse, Misrak Makonnen, Addis Tamire, Helina Worku, Mesele Damte Argaw

## Abstract

**Introduction:** Lack of access to essential health services has remained a significant public health challenge in sub-Saharan Africa. The conventional health service delivery models often fail to account for the mobility patterns and socio-cultural values of these communities. These communities are frequently affected by recurrent droughts, conflicts, floods, and disease outbreaks. This study assessed the contribution of a mobile health services (MHS) delivery model designed to improve access to and utilisation of maternal, neonatal, and child health services in pastoralist contexts of Ethiopia.

**Methods:** A multi-method embedded implementation research with a non-randomized controlled quasi-experimental study design was employed in four selected Woredas of Ethiopia. Baseline data from July 2022 to June 2023 and endline data from July 2023 to June 2024 were extracted from DHIS2. The MHS delivery model was co-designed with stakeholders, redefined community-centred outreach sites, and established a multidisciplinary team, that was implemented monthly. The average distance between the health facility and the centre of the targeted community village was 29.6 kilometres. Data were captured from routine health service records, MHS databases, and program reports, and 143 in-depth interviews among service providers and beneficiaries. Quantitative data were analysed using descriptive statistical methods and difference-in-differences using Stata 18, while qualitative data were coded and analysed using ATLAS.ti.

**Results:** During the twelve months, the MHS reached 79,404 people through 190 sessions held and offered an integrated essential health service in 20 hard-to-reach villages. In the study area, ANC use was 64.0% in Chifra and 75.6% in Dasenech. In addition, penta-3 antigen vaccination improved from 66.0% to 69.7% in Chifra and from 91.2% to 102.1% in Dasenech, respectively. In the difference-in-differences analysis, the MHS significantly enhanced access to ANC follow-up and completion of pentavalent and full vaccination coverage. A Woreda-led approach, community engagement, context-tailored and timely communication, and comprehensive service delivery by a multidisciplinary team enhanced the use and acceptability of the MHS in the pastoral setting. The contribution of MHS to ensuring access and use of MNCH services ranged from 13.0% to 19.1% for continuing family planning and antenatal care, respectively.

**Conclusions:** The study highlights that the MHS delivery model is a suitable platform that has substantially contributed to improving access to and use of maternal, newborn, and child health services in pastoral, agropastoral, and scattered settlements. Being a woreda-led, participatory co-design, community-centred approach, and context-tailored health service delivery approach enhanced the acceptability and effectiveness of the MHS in reaching the underserved community. Integration of MHS with the existing PHC system, with an innovative, sustainable financing mechanism, is recommended to ensure access and use of MNCH in hard-to-reach settings.

## Introduction

Poor maternal, newborn, and child healthcare outcomes remain a significant public health problem in low- and middle-income countries (LMICs) (1). Access to facility-based health services is very limited in pastoral settings, with more than half (58%) of the community in pastoral settings lacking access to essential health services, and less than one-third of them have awareness about immunisation (2). Hence, conventional health service packages and arrangements do not usually consider the mobility pattern, climate crisis, drought, disease outbreak, natural disaster, conflict, and socio-cultural values of pastoral communities (3). In sub-Saharan Africa, more than 50 million inhabitants reside and depend on pastoralism (4), and 20 million (40%) of pastoralist people live in five Horn of Africa countries (5), which have significant challenges in accessing health services. Similarly, in Ethiopia’s pastoral and semi-pastoral settings, residents have struggled with climate change, consecutive seasons of drought, natural disasters, disease outbreaks, and recurrent conflicts (6,7), which disrupted health services, displaced thousands of people, making them more vulnerable to life-threatening conditions (8). In addition, in a stable situation, due to mobility and settlement patterns, low health-seeking behaviour, physical inaccessibility, and gender-related cultural norms, only one-third of the pastoralist communities are potentially accessible to health services within five kilometres of distance to the nearest health facility (9).

The existing service delivery modalities, including integrated periodic outreach service, OneHealth approach, satellite health facilities, periodic campaigns, are standalone approaches with limited scale testing, implemented in designated areas, and not well integrated into the woreda PHC system with limited local government ownership. The woreda-led, PHC level multidisciplinary team-based MHS delivery model is integrated with the routine health system work stream and is a sustainable service delivery model for contexts with low density of health professionals, providing health services to scattered settlements and far distances between communities and health facilities (10). The Mobile Health and Nutrition Team (MHNT) service delivery model has delivered preventive, promotive, and curative healthcare services for underserved, conflict- and drought-affected communities in Africa and Asia (11–13). This service was offered to pastoral communities and internally displaced people in response to emergencies as an alternative model to mitigate crises in several African and Asian countries (14). A series of evidence has indicated that the MHNT service delivery model has significantly improved access to and use of essential health services among hard-to-reach and underserved communities (6,15,16).

In 2004, the Government of Ethiopia (GoE), in partnership with donors and development partners, adapted a tailored multifaceted community-based MHNT Service delivery model (17) and implemented it in humanitarian settings (15,16,18). The MHNT has played a critical role in providing lifesaving health services such as screening for malnutrition, providing routine immunisation services, antenatal care, and medical consultations (9,16,18). Despite this effort, the MHNT service has not been optimally integrated into the health system as a sustainable health service delivery model to address equity for unreached communities (11,16,19,20). In addition, the Ministry of Health (MOH) has developed an implementation guideline (17) to integrate the MHS delivery model in the routine health system in 2023 (21,22). Since April 2022, the Improve Primary Health Care Service Delivery (IPHCSD) project, an investment of the Gates Foundation, implemented by Amref Health Africa and JSI, has tested the MHS strategy to improve access to equitable quality RMNHC-N services through a standardised and context-tailored delivery model (17). Therefore, we aimed to design package of intervention and test using the Integrated Mobile Health Service (IMHS) delivery model to develop a prototype for scale-up and improving access to and utilisation of maternal, neonatal, and child health services in pastoralist communities. Hence, continuous technical support was given in the institutionalisation of sustainable MHS as a routine health service delivery model. This study assessed the implementation of the integrated MHS delivery model to improve maternal, neonatal, and child health service coverage in the pastoralist regions of Ethiopia.

## Methods

### Ethical consideration

The research protocol of this multimethod quasi-experimental study design was reviewed and approved by the Ethiopian Public Health Association (EPHA) research ethical review board (Ref: EPHA/OG/154/24). The study was conducted in line with the recommendations and principles of the Declaration of Helsinki. Informed verbal and written consent were obtained from each study participant voluntarily to be included in the study. Permission was sought to use the anonymous project data from Amref Health Africa, Chifra, Dasenech, Telalek, and Hammer Woreda Health Offices. The collected data was kept confidential and stored in a protected computer after ensuring anonymity through the de-identification of names and other personal identifiers from the record/sheet. Parents/guardians have provided written informed consent for minor study participants and legally authorized representatives for illiterate participants.

### Study setting and context

Ethiopia is the second most populous country in Africa, next to Nigeria. Approximately one-tenth (14 million) of its people live in the lowland areas of Somali, Afar, Oromia, and South Ethiopia regions. In addition, more than two-thirds of its pastoralist population have no access to static facility-based health services (2,21). Pastoralist people depend on livestock, where people seasonally move with their cattle searching for pasture lands and water. Furthermore, the pastoralist population is vulnerable to recurrent outbreaks, conflicts, droughts, and famine. The MHS delivery is a designed strategy to deliver essential health services included in the HEP Optimisation Roadmap 2020-2035 (21), through an integrated innovative PHC service delivery platform in the health system in the agrarian, pastoral, and remote agropastoral settings of Ethiopia. The MHS delivery model was implemented in two woredas, namely Chifra in Afar and Dasenech in the South Ethiopia region. In Chifra woreda, 10 villages (MHS sites) with a total population of 27,739 were identified as hard-to-reach communities that are eligible for MHS. The average distance of these villages (MHS sites) from their respective catchment health centres was 36.6 kilometres. Similarly, in Dasenech woreda, 10 MHS sites with a population of 51,665 were identified as vulnerable and eligible for MHS. The average distance of a village to the nearest health centre was 29.6 kilometres (**Table 1**). The non-intervention woredas, Telalek from Afar and Hammer from the South Ethiopia were selected considering their geographical proximity, similar living arrangements, including culture, norms, values, mobility, seasonal variations, and population livelihood.

**Table 1:**
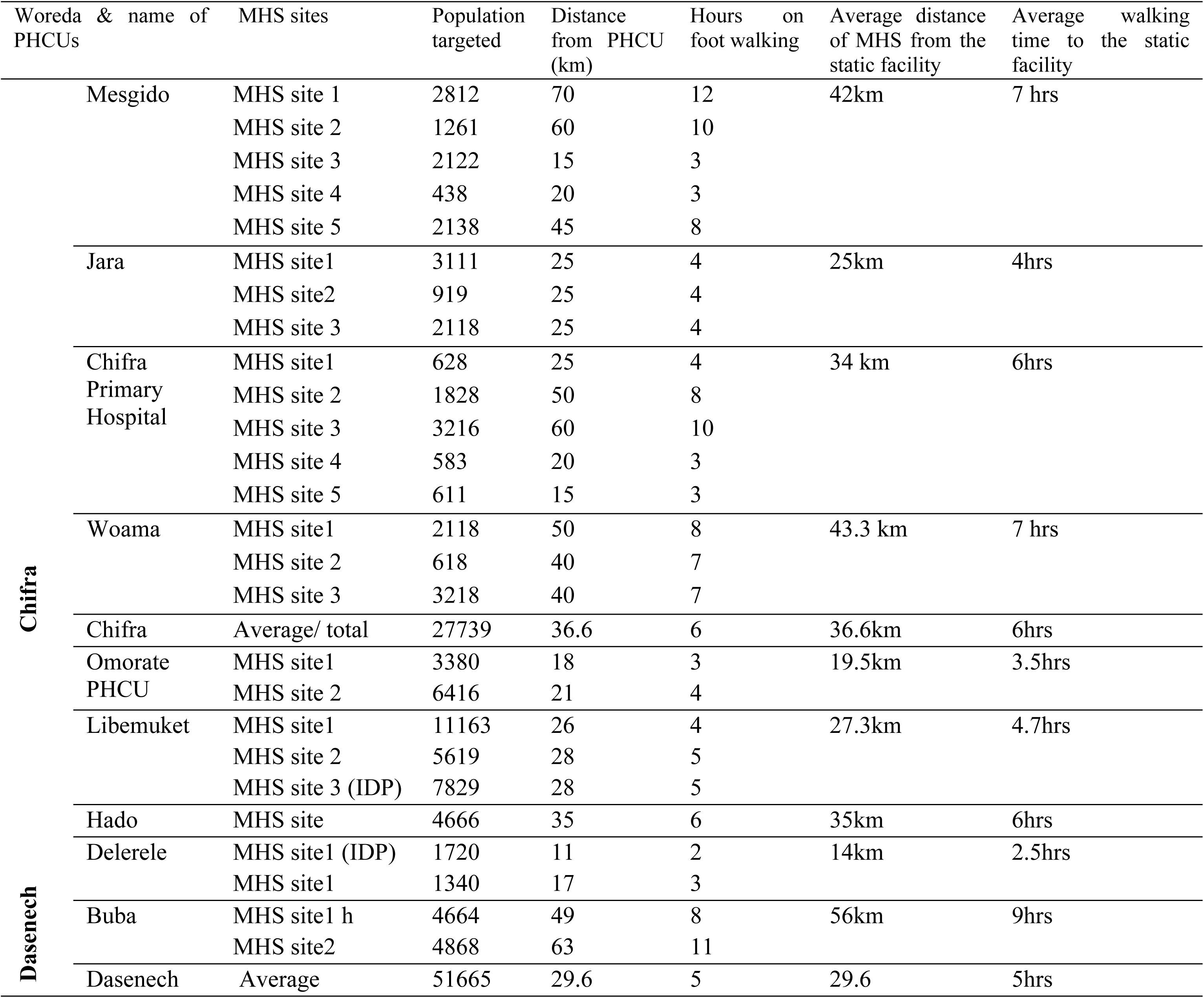
Profile of health facilities and villages targeted for MHS 2023-2024.

| Woreda & name of PHCUs |  | MHS sites | Population targeted | Distance from PHCU (km) | Hours on foot walking | Average distance of MHS from the static facility | Average walking time to the static facility |
|---|---|---|---|---|---|---|---|
| **Chifra** | Mesgido | MHS site 1 | 2812 | 70 | 12 | 42km | 7 hrs |
|  |  | MHS site 2 | 1261 | 60 | 10 |  |  |
|  |  | MHS site 3 | 2122 | 15 | 3 |  |  |
|  |  | MHS site 4 | 438 | 20 | 3 |  |  |
|  |  | MHS site 5 | 2138 | 45 | 8 |  |  |
|  | Jara | MHS site1 | 3111 | 25 | 4 | 25km | 4hrs |
|  |  | MHS site2 | 919 | 25 | 4 |  |  |
|  |  | MHS site 3 | 2118 | 25 | 4 |  |  |
|  | Chifra Primary Hospital | MHS site1 | 628 | 25 | 4 | 34 km | 6hrs |
|  |  | MHS site 2 | 1828 | 50 | 8 |  |  |
|  |  | MHS site 3 | 3216 | 60 | 10 |  |  |
|  |  | MHS site 4 | 583 | 20 | 3 |  |  |
|  |  | MHS site 5 | 611 | 15 | 3 |  |  |
|  | Woama | MHS site1 | 2118 | 50 | 8 | 43.3 km | 7 hrs |
|  |  | MHS site 2 | 618 | 40 | 7 |  |  |
|  |  | MHS site 3 | 3218 | 40 | 7 |  |  |
|  | Chifra | Average/ total | 27739 | 36.6 | 6 | 36.6km | 6hrs |
| **Dasenech** | Omorate PHCU | MHS site1 | 3380 | 18 | 3 | 19.5km | 3.5hrs |
|  |  | MHS site 2 | 6416 | 21 | 4 |  |  |
|  | Libemuket | MHS site1 | 11163 | 26 | 4 | 27.3km | 4.7hrs |
|  |  | MHS site 2 | 5619 | 28 | 5 |  |  |
|  |  | MHS site 3 (IDP) | 7829 | 28 | 5 |  |  |
|  | Hado | MHS site | 4666 | 35 | 6 | 35km | 6hrs |
|  | Delerele | MHS site1 (IDP) | 1720 | 11 | 2 | 14km | 2.5hrs |
|  |  | MHS site1 | 1340 | 17 | 3 |  |  |
|  | Buba | MHS site1 h | 4664 | 49 | 8 | 56km | 9hrs |
|  |  | MHS site2 | 4868 | 63 | 11 |  |  |
|  | Dasenech | Average | 51665 | 29.6 | 5 | 29.6 | 5hrs |

**Table 1.** Key features of the mobile health service.

|  |  |
|---|---|
| Service package | The essential health services delivered through district-led integrated MHS, include: Under five children health service, Immunization; Family planning; ANC & PNC services, and screening for malnutrition; Adult patient diagnosis and treatment and other (HIV testing and awareness, TB screening & awareness, Hygiene and sanitation, referral) |
| Team composition | A multidisciplinary team established and deployed on regular schedules which consist Health officers/BSc nurses, Laboratory professionals, Pharmacy professionals, Health extension workers and community voulunters, called Village Health Leaders (VHL) |
| Community Mobilization | The frontline community helath workforce cadres, known as HEWs and community volunteers, called VHLs communicate the mobile health service site and schedule and leverage the existing community-level communication tradition (eg.Dagu in Afar) |
| Governance | Coordinated by the woreda health office whp avail necessary logistics and fleet management in collaboration with relevant stakeholders; including availing vehicle, medicines and supplies; HMIS tools used for recording and reporting; carry out regular Supportive Supervision and Performance review and management and other technical and logistical support |
| Data and reporting | The multidisciplinary team used paper-based HMIS recording tool to document the MHS services at field and entered into DHIS2, however, the registration has not reflected the MHS services contribution. |

### Study design

A quasi-experimental study was conducted using a multi-method embedded implementation research approach. The IPHCSD project employed an iterative co-design process involving diverse stakeholders to develop and support a woreda-led Mobile Health Service (MHS) strategy. The strategy was implemented through multidisciplinary teams and included the definition of service packages and delivery mechanisms, with active participation from health professionals, community leaders, and community members to ensure contextual relevance, ownership, and sustainability (Fig 1). The implementation of the IPHCSD project strategies was guided by the Reach Effectiveness Adoption Implementation and Maintenance (RE-AIM) framework (23), and an embedded implementation research (EIR) (24) approach with a hybrid quasi-experimental design (25,26) was conducted using multimethod data sources from July 2022 to June 2023 for baseline and July 2023- June 2024 for endline. Overall learning documentation of MHS was written following the Standard Reporting Items for Implementation Research (27) and the consolidated reporting format for interventional studies (28).

**Fig 1.**
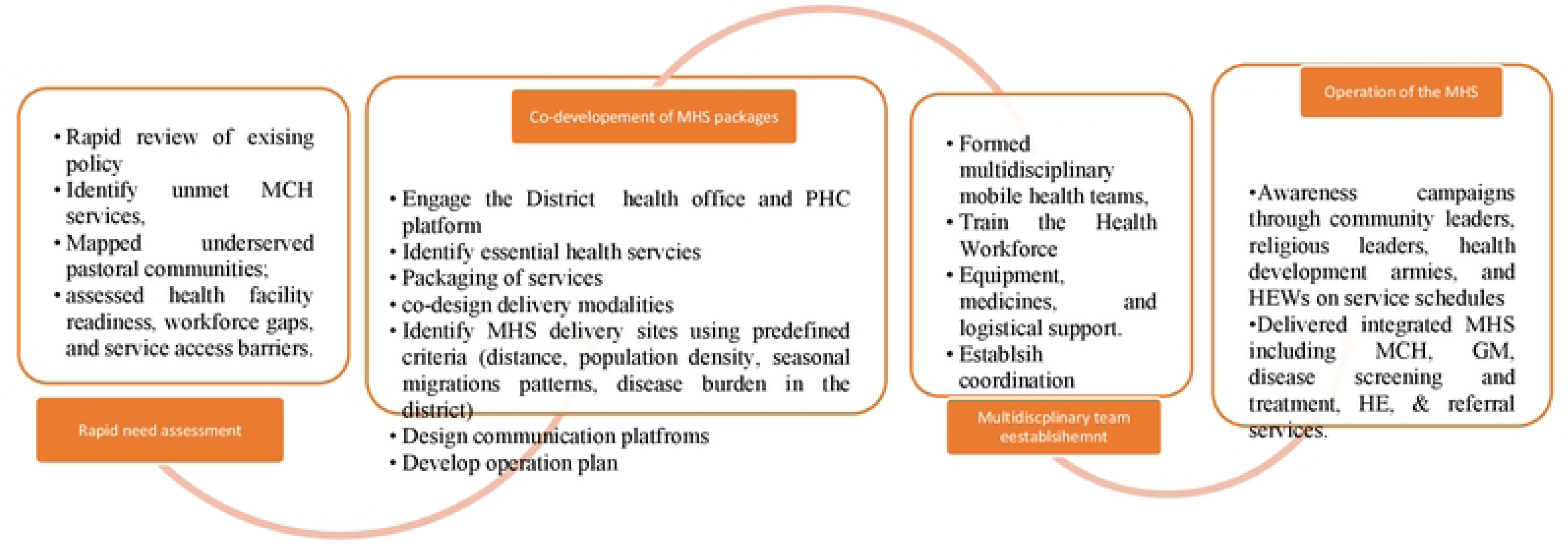
Iterative pathway for the MHS delivery strategy co-development and implementation

### Intervention co-development and description

The IPHCSD project conducted a rapid assessment to identify the health needs of pastoralist communities and document lessons learned from MHNT deployed to address the emergency health and nutrition crisis.

In addition, the participants identified 20 hard-to-reach sites based on the population density, accessibility, and health service needs of pastoralist communities. The woreda health office established a coordinating and multidisciplinary team comprising health officers, nurses, midwives, health extension workers, community volunteers, and drivers. The coordination team collects drugs, vaccines, fuel, supplies, tents, furniture, and medical equipment. Each team received a comprehensive orientation on culturally sensitive MHS packages and reporting requirements. The MHTs have offered a day-long essential health service every month in 20 sites. The IPHCSD project has also supported each woreda to conduct supportive supervision, organise learning sessions, and review meetings to track progress and performance of MHSs.

### Sampling methods and data sources

The four woredas were purposefully selected based on similar characteristics in living arrangement, geographic proximity, and cultural values. At the time of this assessment, the MHS had been implemented for twelve months in two woredas. Multimethod data collection approaches were used to gather comprehensive data from routine records of health facilities, intervention implementation, regular performance reviews, and process evaluation reports. MHS beneficiaries’ data were captured using the DHIS2 platform, which was designed for the IPHCSD project. Additionally, key informants were purposively recruited from service beneficiaries, community representatives, Village Health Leaders, Healthcare Providers, Woreda Health Offices, and implementing partners. Lessons learned, review meeting, and process evaluation reports were reviewed. The data of MHSs were captured using routine health information systems registers, tally sheets, and reporting forms developed for health posts and health centres in Ethiopia. The data were extracted using semi-structured and structured tools developed in Microsoft Excel. In addition, socio-demographic characteristics of service beneficiaries were captured from MHS documents. Diagnostic, curative, health promotion, and rehabilitation information were captured using pretested tools. In addition, relevant stakeholders and beneficiaries were interviewed using a semi-structured interview guide to explore availability, accessibility, affordability, adequacy, and acceptability of MHS (29). The face-to-face interviews last 45-60 minutes. High-level experts’ insights, feedback, and recommendations were gathered during the provision of technical assistance, monitoring of project implementation, supportive supervision, and performance review meetings. Additionally, evaluation of the program implementation was conducted and details of the interview guide (S1 interview guide).

### Measurement of outcome

The contribution of MHS is depicted for selected MCH services indicators, which were computed from routine services records and using the estimated Woreda target population as a denominator. Antenatal care, institutional delivery, postnatal care, contraceptive acceptance rate (CAR), and new family planning services utilization, nutritional screening, provision of nutrition supplements, and acute childhood illness management were used to measure MHS outcomes. Eleven maternal and child health key indicators were identified and used to estimate the effect of the IPHCSD project MHS support on targeted program outcomes. The indicators used to measure maternal health outcomes include Contraceptive Acceptance Rate (CAR), the first Antenatal Care (ANC1), the ANC4, and Postnatal Care (PNC) coverage. In addition, the key indicators used to measure child health outcomes consist of the first Pentavalent vaccination (Penta 1), the third Pentavalent vaccination (Penta 3), the first Measles Containing Vaccine (MCV1), the Second Measles Containing Vaccine (MCV2), Full Immunization, treatment of pneumonia, and diarrhea for children. The contribution of the MHS in improving access to MCH services was assessed using the 5As domain for health care access (29) embedded with the RE-AIM framework (23) (**Table 2**).

**Table 2.**
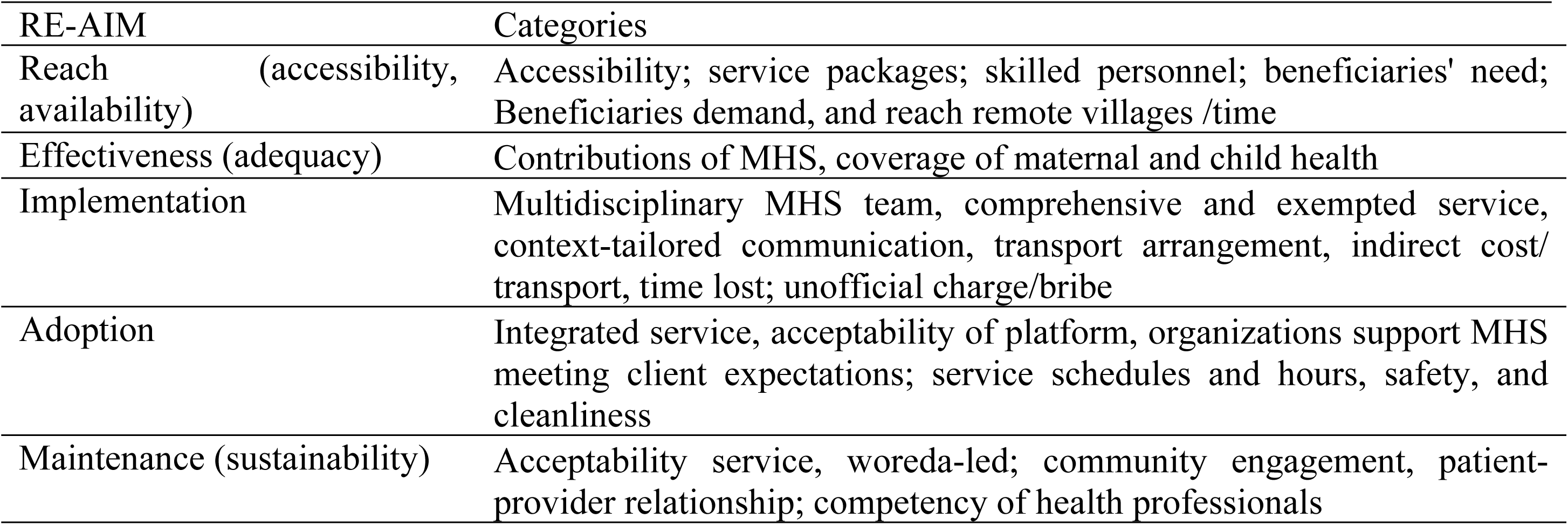
Thematization of MHS contribution to MCH services.

### Data management and analysis

Data cleaning and analysis were done with Microsoft Excel Office 2016 and Stata 18 (S2 data set). A descriptive analysis using frequency tables and graphs was used to present the findings of categorical data. The contribution of MHSs to child health and maternal health service utilisation was determined using proportions of twelve months of data (i.e., July 2023 – June 2024). The analysis described the overall diagnostic, curative, immunisation, antenatal care, postnatal care, family planning and referral service beneficiaries by woreda was employed to check the presence of statistical differences in health service coverage over consecutive months. In addition, a test of two proportions was employed to determine differences between six rounds of mobile health services. Difference-in-Differences (DID) analysis was computed, and a statistical association was declared at a *p-*value of less than 0.05. Qualitative data analysis was done using a content analysis approach focusing on availability, accessibility, affordability, adequacy, and acceptability parameters of the access to health care analysis framework (29). The collected data, using handwritten notes and interview records, were transcribed, coded using ATLAS.ti, and arranged thematically (30). Data triangulation was used to verify and validate quantitative results.

## Results

### Characteristics of study participants

More than half (61.5%) of the qualitative interview respondents were males, 72.7% were currently married, and the highest number of respondents (38.5%) were nurses. Among 46 HEWs, 80.4% have a level-IV certificate, and nearly all (88.8%) were either head of the facility or a team lead in the facility. 65.7% of respondents live within the village (**Table 3**).

**Table 3.** Distribution and background characteristics of HWs surveyed about MHS strategy (n=143)

| **Variables** | **Categories** | **N** | **%** |
|---|---|---|---|
| Sex | Female | 55 | 38.46 |
|  | Male | 88 | 61.54 |
| Level of Education | College certificate | 76 | 53.15 |
|  | First Degree | 57 | 39.86 |
|  | Second Degree and above | 8 | 5.59 |
|  | Other | 2 | 1.40 |
| Profession | Health Extension Worker | 46 | 32.17 |
|  | Nurse | 55 | 38.46 |
|  | Midwife | 16 | 11.19 |
|  | Health Officer | 17 | 11.89 |
|  | Environmental Health | 1 | 0.70 |
|  | Medical Lab | 2 | 1.40 |
|  | Pharmacist/Druggist | 1 | 0.70 |
|  | Other | 5 | 3.50 |
| Position in the health facility | Head of the health facility | 59 | 41.26 |
|  | Focal person | 68 | 47.55 |
|  | Staff with no managerial role | 16 | 11.19 |

In both intervention and non-intervention woredas, 23% of the estimated population were women of reproductive age (15-49 years), 2.8% were pregnant mothers, 2.6% were surviving infants, and 11.4% were under-five children (**Table 4**).

**Table 4.**
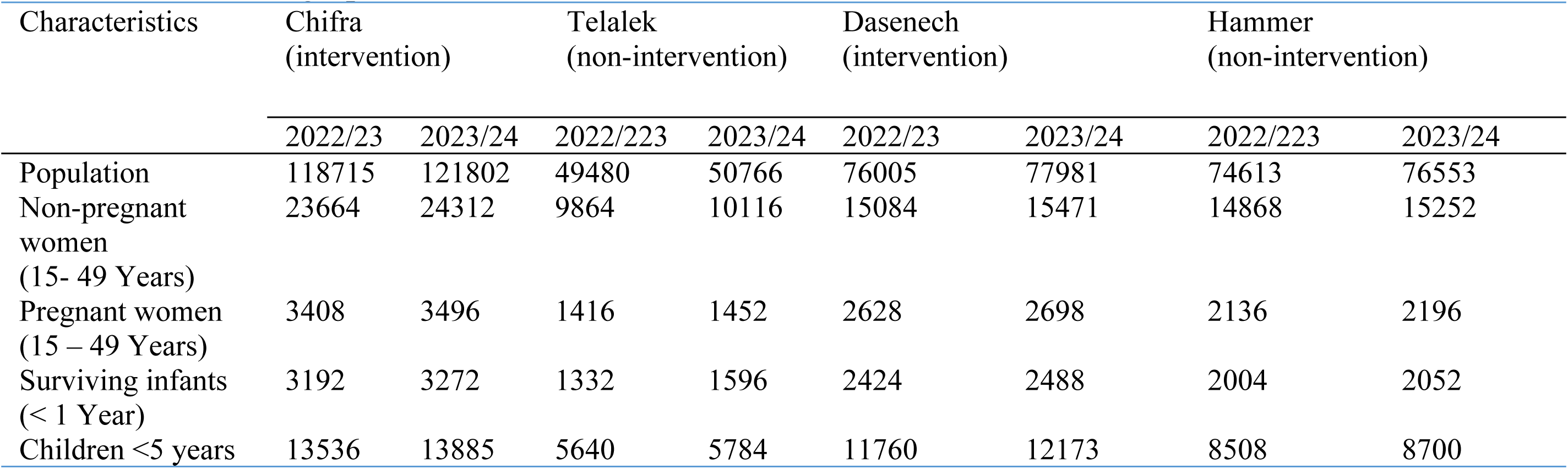
Socio-demographic characteristics of intervention and non-intervention.

| Characteristics | Chifra<br>(intervention) |  | Telalek<br>(non-intervention) |  | Dasenech<br>(intervention) |  | Hammer<br>(non-intervention) |  |
|---|---|---|---|---|---|---|---|---|
|  | 2022/23 | 2023/24 | 2022/223 | 2023/24 | 2022/23 | 2023/24 | 2022/223 | 2023/24 |
| Population | 118715 | 121802 | 49480 | 50766 | 76005 | 77981 | 74613 | 76553 |
| Non-pregnant women<br>(15- 49 Years) | 23664 | 24312 | 9864 | 10116 | 15084 | 15471 | 14868 | 15252 |
| Pregnant women<br>(15 – 49 Years) | 3408 | 3496 | 1416 | 1452 | 2628 | 2698 | 2136 | 2196 |
| Surviving infants<br>(< 1 Year) | 3192 | 3272 | 1332 | 1596 | 2424 | 2488 | 2004 | 2052 |
| Children <5 years | 13536 | 13885 | 5640 | 5784 | 11760 | 12173 | 8508 | 8700 |

### Reach and adoption of mobile health services

In this study, maternal, neonatal, and child health and adult outpatient health services were packaged to meet the needs and demands of underserved or marginalised communities in both intervention woredas. During the twelve months, 79,404 people were served through organising 190 service sessions in 20 villages. The majority of respondents listed eight health services or program packages which were offered at MHS sites, namely, health promotion and education, antenatal care, postnatal care, family planning, nutritional screening, immunisation, deworming, adult and under-five outpatient services.

The MHS delivery model is integrated into the routine health system in mobilising a multidisciplinary team, drugs, supplies, and logistics from the nearby catchment health facility.

> *“The health facility conducted a rapid assessment, identified the most common health problems of the community, and the MHS sites. Then, a multidisciplinary health professional was deployed and offered transport services with drugs and supplies. In addition, development partners have provided medical equipment, office furniture, and emergency drug kits. Furthermore, the cost of fuel and daily subsistence allowances of health professionals were covered by Amref Health Africa.”* [WorHO, KII-35].

In addition, key informants expressed that in their Woredas, a large number of women and children may not have access to essential health services unless the MHS delivery model were practiced in the pastoral settings. A key informant remarked that:

> *“…pregnant women and children received ANC, immunization, outpatient, and other lifesaving services at MHS sites. The MHS is the main platform in ensuring availability of essential health care for hard-to-reach communities”* [KII, F, 103]

### Effectiveness of MHS

Chifra and Dasenech woredas are MHS intervention (I) sites, while Telalek and Hammer woredas are non-intervention (NI) sites of the IPHCSD project. The demographic characteristics of targeted woredas. In the year 2022/23, the projected population was 11 8,715 and 76,005 for the intervention woreda, i.e., Chifra and Dasenech, respectively. In comparison, the non-intervention woredas have 49,480 and 74,613 people for Telalek and Hammer woredas, respectively. The baseline data from July 2022 to June 2023 was compared to the endline (implementation period) data from July 2023 to June 2024. In Chifra woreda, 1,677 women in the reproductive age group have received family planning service as new acceptors. During the same period, only 32 women (1.9%) received FP services at ten MHS sites. In Dasenech woreda, 15,981 non-pregnant women in the reproductive age group received FP services at all service points, of whom 1,681 (10.5%) of the women were served at ten MHS sites. In addition, 2454 and 4082 pregnant mothers received their first antenatal care services at all health service delivery sites in Chifra woreda. Out of these pregnant mothers, 568 (23.1%) in Chifra and 637 (15.6%) in Dasenech were served at MHS sites in Chifra and Dasenech woredas, respectively. In Chifra Woreda, 2480 infants received first pentavalent (Penta 1) vaccination. Of these, 1,074 (43.3%) were vaccinated at MHS sites. Similarly, 2,597 infants under one year old were vaccinated in Dasenech woreda (**Fig 2**). Of these, 602 (23.2%) infants were vaccinated at ten MHS sites. Overall, MHS contributed one-third (704; 36.5%) and one-fifth (545; 22.3%) of complete immunisation services in Chifra and Dasenech woredas, respectively (**Table 5**).

**Fig 2.**
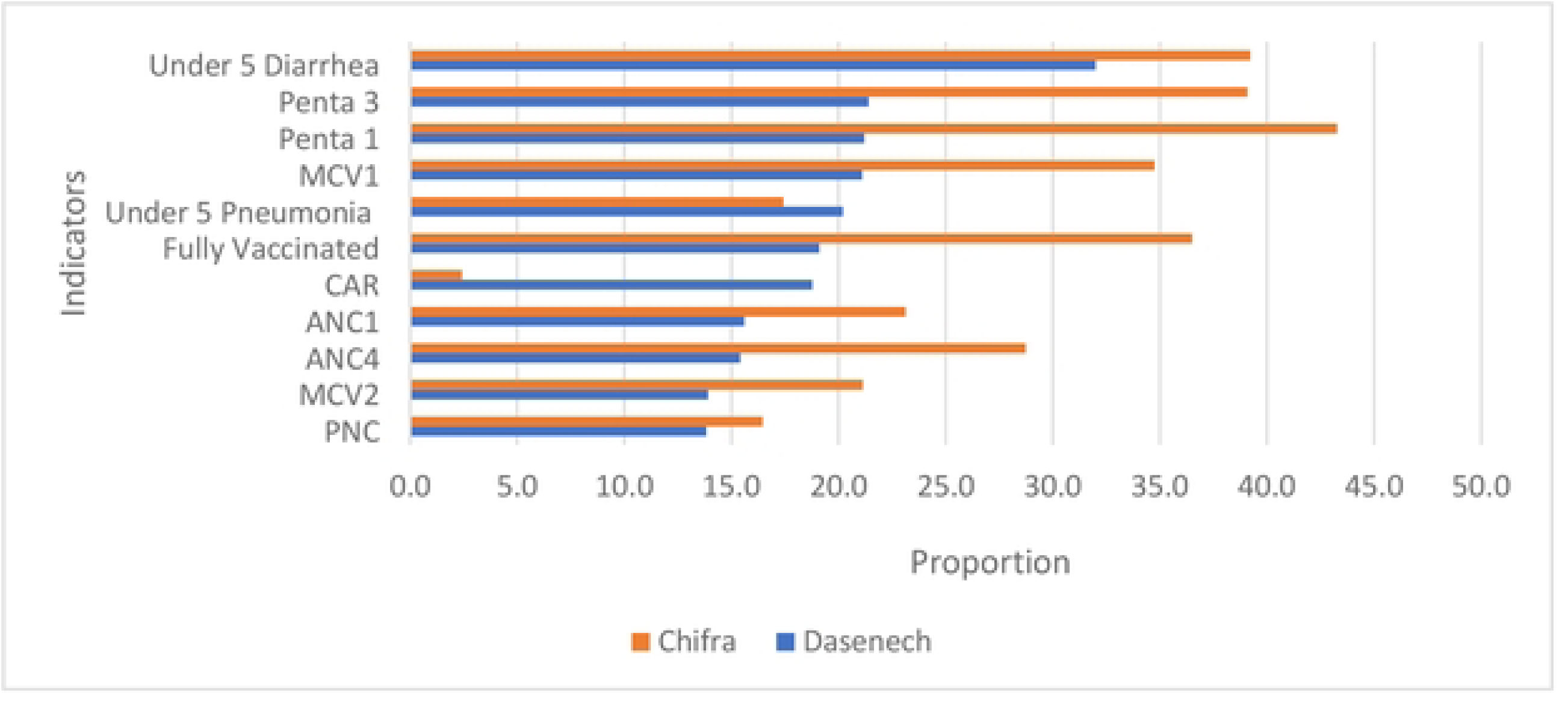
The contribution of integrated MHS delivery modality for the pastoral community in Ethiopia

**Table 5.** Contribution of MHS on maternal and child health services coverage in interventions woredas, July 2023 - June 2024.

| MHS contributions | Chifra |  |  | Dasenech |  |  |
|---|---|---|---|---|---|---|
|  | All service points | MHSs | Contribution (%) | All service points | MHSs | Contribution (%) |
| CAR | 1677 | 32 | 1.9 | 15981 | 1681 | 10.5 |
| ANC1 | 2454 | 568 | 23.1 | 4082 | 637 | 15.6 |
| ANC4 | 1290 | 371 | 28.8 | 3410 | 526 | 15.4 |
| PNC | 1024 | 166 | 16.2 | 2814 | 389 | 13.8 |
| Penta 1 | 2480 | 1074 | 43.3 | 2597 | 602 | 23.2 |
| Penta 3 | 2280 | 888 | 38.9 | 2547 | 575 | 22.6 |
| MCV1 | 2110 | 734 | 34.8 | 2480 | 611 | 24.6 |
| MCV2 | 1698 | 359 | 21.1 | 2448 | 554 | 22.6 |
| Fully Vaccinated | 1927 | 704 | 36.5 | 2443 | 545 | 22.3 |
| Under 5 Pneumonia Treatment | 1208 | 212 | 17.5 | 2142 | 521 | 24.2 |
| Under 5 Diarrhea Treatment | 1846 | 726 | 39.3 | 3116 | 1384 | 44.4 |

### Maternal Health Service coverages

The PNC coverage rose from 24.5% at baseline to 29.3% at the endline, showing an increase in coverage by 4.8 percentage points in Chifra. While in Telalek, a non-intervention woreda, PNC coverage decreased from 34.1% at baseline to 20.8% at the endline. Hence, the net difference of 8.5 percentage points is statistically significant at a 95% confidence level. Similarly, in Dasenech, another intervention woredas which has a PNC coverage of 99.6 % and 104.3% at baseline and endline evaluation. The MHS interventions increased the PNC service by 4.7 percentage points. The PNC coverage in Hammer Woreda (comparison site) was 78.0% and 110.6% at baseline and endline assessment, respectively. The net difference was negative 27.9 percentage points, which is statistically significant at a 95% confidence level (Table 4).

### Child health service coverages

The first dose of the pentavalent vaccine coverage rose from 72.1% at baseline to 75.8% at the endline in Chifra, revealing an increased service coverage by 3.7 percentage points. While in Telalek woreda, the Penta-1 vaccination coverage has decreased by 17.7 percentage points, i.e., from 99.1% to 81.4% of service coverage. Therefore, the net difference of 21.4 percentage points is a statistically significant difference at a 95% confidence level. Similarly, in Dasenech woreda, first-dose pentavalent vaccination coverage has increased from 93.5% at baseline to 104.4% at endline in the intervention woreda. While in Hammer woreda, the first dose of pentavalent vaccine coverage increased from 129.9% at baseline to 132.7% at the endline evaluation. The net difference of 8.1 percentage points is not statistically significant at a 95% confidence level (Table 6).

**Table 6.** The effect of MHS on maternal and child health services use from July 2022 to June 2024.

| Indicators | Baseline |  |  |  |  | End line |  |  |  |  | DID 95%CI |
|---|---|---|---|---|---|---|---|---|---|---|---|
|  | Intervention | % | Comparison | % | Diff | Intervention | % | Comparison | % | Diff |  |
| Chifra Vs Telalek |  |  |  |  |  |  |  |  |  |  |  |
| CAR | 2532 | 10.7 | 2426 | 24.6 | -13.9 | 1677 | 6.9 | 2023 | 20.0 | -13.1 | 0.8 (-2.22,3.04) |
| ANC1 | 2065 | 60.6 | 1336 | 94.4 | -33.8 | 2454 | 70.2 | 1193 | 82.2 | -12 | 21.8(12.51,29.09)* |
| ANC4 | 1168 | 34.3 | 659 | 46.6 | -12.3 | 1290 | 36.9 | 644 | 44.4 | -7.5 | 4.8 (-0.99, 9.91) |
| PNC | 834 | 24.5 | 484 | 34.1 | -9.6 | 1024 | 29.3 | 447 | 20.8 | 8.5 | 18.1(12.5, 23.3)* |
| Penta 1 | 2301 | 72.1 | 1320 | 99.1 | -27.0 | 2480 | 75.8 | 1299 | 81.4 | -5.6 | 21.4 (12.7, 28.7)* |
| Penta 3 | 2106 | 66 | 1268 | 95.2 | -29.2 | 2280 | 69.7 | 1237 | 77.5 | -7.8 | 21.4 (12.94,28.06)* |
| MCV1 | 2058 | 64.5 | 1282 | 96.3 | -31.8 | 2110 | 64.5 | 1225 | 76.8 | -12.3 | 19.5 (12.7, 30.1)* |
| MCV2 | 1592 | 49.9 | 1193 | 89.6 | -39.7 | 1698 | 51.9 | 1040 | 65.2 | -13.3 | 26.4 (10.5, 26.9)* |
| Fully Vaccinated | 1669 | 52.3 | 1173 | 88.1 | -35.8 | 1927 | 58.9 | 1154 | 72.3 | -13.4 | 22.4 (15.8, 34.80* |
| Under 5 Pneumonia | 1191 | 8.8 | 2143 | 38.0 | -29.2 | 1208 | 8.7 | 1440 | 24.9 | -16.2 | 13.0 ( 7.71,16.69)* |
| Under-5 diarrhea | 1705 | 12.6 | 1178 | 20.9 | -8.3 | 1846 | 13.3 | 1006 | 17.4 | -4.1 | 4.2 (0.7, 7.3) |
| Dasenech Vs Hammer |  |  |  |  |  |  |  |  |  |  |  |
| CAR | 13424 | 89 | 8712 | 58.6 | 30.4 | 15981 | 103.3 | 10600 | 69.5 | 33.8 | 3.4 (-16.5, 23.1) |
| ANC1 | 3392 | 129.1 | 3080 | 144.2 | -15.1 | 4082 | 151.3 | 3335 | 151.9 | -0.6 | 14.5 (-7.7, 36.6) |
| ANC4 | 2885 | 109.8 | 1916 | 89.7 | 20.1 | 3410 | 126.4 | 2518 | 114.7 | 11.7 | -8.4 (-30.7, 14.0) |
| PNC | 2617 | 99.6 | 1666 | 78.0 | 21.6 | 2814 | 104.3 | 2428 | 110.6 | -6.3 | -27.9 (-49.4,-6.4)* |
| Penta 1 | 2266 | 93.5 | 2603 | 129.9 | -36.4 | 2597 | 104.4 | 2723 | 132.7 | -28.3 | 8.1(-7.9,24.3) |
| Penta 3 | 2220 | 91.6 | 2180 | 108.8 | -17.2 | 2547 | 102.4 | 2538 | 123.7 | -21.3 | -4.1(-18.0,9.6) |
| MCV1 | 2111 | 87.1 | 2064 | 103.0 | -15.9 | 2480 | 99.7 | 2439 | 118.9 | -19.2 | -3.3(-20.9,14.0) |
| MCV2 | 3046 | 125.7 | 1829 | 91.3 | 34.4 | 2448 | 98.4 | 2472 | 120.5 | -22.1 | -56.5(-148.1,35.2) |
| Fully Vaccinated | 2208 | 91.1 | 2020 | 100.8 | -9.7 | 2443 | 98.2 | 2380 | 116.0 | -17.8 | -8.1(-29.3,12.9) |
| Under 5 Pneumonia | 1846 | 15.7 | 1097 | 12.9 | 2.8 | 2142 | 17.6 | 1052 | 12.1 | 5.5 | 2.7(-4.6,10.1) |
| Under-5 diarrhea | 4421 | 37.6 | 2246 | 26.4 | 11.2 | 3116 | 25.6 | 2705 | 31.1 | -5.5 | -6.7(-16.0,2.7) |

*NB.: \* P <0.05; I: Intervention; NI: None Intervention; CAR: Contraceptive Acceptance Rate; ANC: Antenatal Care; PNC- postnatal care; MCV- Measles containing vaccine; Diff- difference in proportions; DID- Difference in Difference; CI- Confidence Interval*

### Accessibility of mobile health service

This theme is described with the average distance of an MHS site from a health centre or health posts. In addition, the average walking hours from the centre of a village to reach the nearest health centre or health posts were presented (**Table 1**). The list of villages with average population, distance in kilometres, and walking time in hours by primary health care unit and woreda is presented below.

> *“…considering Afar communities’ mobility, the MHS are arranged with prior communication about the site centered on the community. The MHS is provided for hard-to-reach areas, people who have difficulties in reaching static health facilities. Most of the facilities are 15 to 20 km from MHS sites.” [*KII, 01]

The mobile health service has increased the potential health service coverage. The MHT has been travelling 30 km and providing various services, including ANC, outpatient consultations, nutritional screening, and immunisation. A participant from Dasenech mentioned health service access before MHT as follows:

> *“Previously, the services were provided only in fixed health facilities. There was only an outreach program for EPI, ANC, and FP, but as you know, the outreach program covers a radius of 5 kilometres or less. Still, now you need to travel more than 30 km and provide service for the hard-to-reach areas where the community moves seasonally, in which functionalizing MHS is not an easy task.”* [DPP, WorHO KII77]

### Implementation fidelity and adaptation

The woreda-led MHS delivery model is comprised of a PHCU-based multidisciplinary MHNT established to serve the catchment population. The MHS sites were selected considering distance and population (Table 1). The MHS is scheduled every month and implemented as per the schedule after conducting community mobilisation by village health leaders. A key informant remarked about the iterative site selection process and program adaptation as follows:

> *“…the sites identified through iterative co-design process that labelled as ‘hard to reach’ and based on our initial assessment/classification as hard to reach areas…in the meantime, WorHO and partners discussed on what type of MHT is needed and what should be included in the packages and which conditions shall be taken into consideration…” [WorHO, KII, MHT coordinator]*

Another KII stated that;

> *“…the MHN team is comprised of nurses, midwives, HO, and HEWs, etc. This team provides the outreach service once a month; however, the number of MHT is not always adequate/ reliable, as it depends on conditions. Most of the time we take three professionals.” [WorHO, KII-15]*

### Stakeholders’ engagement and support

Various stakeholders, like the woreda health office, health facilities, professionals, partners, village administration, community representatives, VHL, and HEWs, have played a key role in establishing and running MHS.

> *“The health facility identified the most common health problems of the community and deployed health professionals from different disciplines. It also offered ambulances, medicines, and medical supplies.”* [MHT-Coordinator, HC, KII-01]
>
> *“The partner, AMREF, supports the woreda with supplies; for instance, examination beds, screens, tables, chairs, fuel for the Ambulance, per diem for health professionals and drivers, all of which are covered by the partner every month.”* [HC/MHT-Coordinator, KII09]

According to the woreda-level collated data from the health facilities assessment, 100% of health facilities out of the total assessed (n=9) had a functional MHNT across two woredas. At the woreda level, co-designing, area selection, and regular updates, role identification for actors, and alignment of the plan of MHS with the PHCU and national strategy were well performed. However, having registration formats and privacy and safety of the setting were reported as low. The performance on service packages was distributed among all expected services in the guideline. However, the professional mix and service site selection were relatively low **(Table 3)**.

A participant also mentioned that before the MHT program, mothers and children were referred to the health facilities after they developed complications. The MHT implementation enabled the community to access care. The participants described it as follows:

> *“…previously, the mothers may arrive at the health facility after developing complications, delayed for two to three days. For instance, during abortion, most of the cases presented at health facilities with severe anaemia or sepsis; some cases may even require referral service. Therefore, before the beginning of the mobile health team, the communities were living in a terrible situation…” [MHN coordinator, HC, KII03]*

The integrated MHS was designed with stakeholders considering the remoteness of the sites, the population, and the centrality of the area to gather the surrounding community. All PHCUs (9, 100%) considered at least one of the suggested criteria to select the community/areas for MHS. The MHS modality targeted to address the dire health problems was under-five children nutritional screening and treatment for acute severe and moderate malnutrition, pneumonia, diarrhoea, malaria, for all, and pregnant women, and FP services were considered by all PHCUs.

> “*We discussed at various meetings about the importance of this approach. Then, we identified the areas labelled as ‘hard to reach’ and based on our initial assessment/classification as hard to reach areas, the team omitted some areas from this category.*” [MHT coordinator KII-22]
>
> “*A rapid assessment has been done at the woreda-level to assess their experience on MHT. Then, the stakeholders discussed with WorHO and designed the type of MHT needed and the service packages to be included in the MHS modality, and which conditions shall be taken into consideration…”* [WorHO, KII04].

### Multidisciplinary mobile health team

The uniqueness of MHS is established by a multidisciplinary team comprised of health professionals, health officers, BSc nurses, laboratory professionals, pharmacy professionals, health extension workers, and volunteer health leaders (VHLs). Furthermore, stakeholders’ roles and responsibilities were outlined, including who would be responsible for covering MHS-related expenditures, fuel, and MHT coordinators were assigned. After the MHT establishment, logistics were arranged, such as tents, chairs, tables, and other commodities. The number of sites selected and assigning the team to specific communities were also major activities in the process of MHS establishment.

> *“In our woreda, there are eight selected sites. The team is comprised of nurse, a midwife, Health Officers, etc. This team provides the MHS once a month…” [HC head, KII17]*

The multidisciplinary MHT has also been supported by the HEWs at health posts and VHLs, ensuring they carry out their duties responsibly.

> “…*the MHT is good at providing the service. However, some have trouble reporting on time. They are somewhat late. Sometimes their incentives become late too…”* [WorHo, K01]

### The MHS sustainability (maintenance)

The MHS delivery model was an integrated and ward-led platform with active community engagement and involvement in arranging the schedule, fostering ownership. In addition, it was a PHC cluster that enhanced local capacity and integration. Acceptability, affordability, and service adequacy are the themes explained in detail below that improve the suitability and sustainability of MHS for the pastoral setting.

### Affordability of mobile health services

The mobile health service (MHS) is being provided free to the community with cost0recovery mechanism by the government and development partners’ support. Mothers, children, elders, and persons with disability who have limited decision-making power to visit health facilities and lack the resources to cover the opportunity cost of utilising fixed health facility services. However, there is no functional community-based health insurance scheme in IPHCSD’s MHS intervention woredas to establish a financial recovery mechanism for MHS services within the health system.

> *“The MHS is being provided free of charge for service beneficiaries. and it has a cost implication on static health centre services through stockout of drugs and supplies.” [HC coord* KII103]

### Adequacy of mobile health services for integration

This theme describes the organization of mobile health services to meet the expectations of service beneficiaries. Client expectations on service schedules, working hours, safety, and cleanliness of mobile health service sites are described below.

The MHS is being provided monthly by an expert leads the MHS team from a woreda health office. As the key informant said:

> “…*it is a comprehensive MHS which addresses the needs of children, mothers, and adult outpatients including immunization, deworming, Vitamin A, Nutrition screening, ANC, PNC, hypertension, and diabetes mellitus screening, and counseling service for GBV survivors.*” [Benefciary-KII-103]
>
> *“…the team is comprised of a nurse, midwife, health officer, health extension worker, village health leaders, and Woreda MHS coordinator. This team provides the outreach service once a month…” [HC head, KII 62]*

The MHS is being implemented once a month, which is not enough to satisfy the mobile or pastoral communities’ needs through a day-long service only.

> *“…the MHS team is scheduled and used to provide health services once a month for a full day in selected MHS sites; however, the community expects and demands more frequent mobile health service provisions.” [HC, IDI01]*

Pregnant women expect to have point-of-care limited obstetric ultrasound services and laboratory investigations.

> *“…pregnant mother demands limited obstetric ultrasound and laboratory services which are available at a primary hospital or a health center.” [ HC,* KII-101]

The MHS team practiced infection prevention and standard precautions for health workers, service beneficiaries, and communities. A health worker has to say:

> *“… we used safety boxes to collect all medical wastes and ensure safe disposal ..*.” [KII, F, 103]

### Acceptability of mobile health services

The Woreda council and community members have a positive perception of the MHS. It addresses the needs of the hard-to-reach communities. In addition, since MHS team members are permanent staff of the nearby health facilities, service beneficiaries have trust in them.

> “…*the community knows the health workers at the health facilities through different engagements, this has created trust in them.*” [KII, M, 101]
>
> *“…such a kind of MHS platform is a model for fostering community trust, addressing the needs and demands of vulnerable communities.”* [KII, M, 101]

The community volunteers (Village Health Leaders, locally named Makafta leaders) in the MHS are taking key roles in coordination.

> *“There are community volunteers who are responsible for informing the communities about the schedule, service packages, and service hours.” [KII, F, 103]*

The community has a good uptake of MHS service.

> “…*in these sites, the communities are satisfied and happy with the MHS, but people who the MHS does not reach have been raising questions. Yet we cannot answer all the needs*.” [KII, M, 101]
>
> *“We are serving them in a very nearby area under a shade in front of their home.” [KII, F, 103]*
>
> *“We have got a lot of benefits from MHT, such as vaccinations, guidance, and nutritional services for someone experiencing malnutrition, such as oatmeal, even though currently their focus is on vaccination…”* [CM, FGD-p02]
>
> *However, the MHS service requires some logistics and supplies to provide client-friendly service.* “…*the privacy of the clients was kept using tents and screens… the community received services without fear of breaching their privacy.”* [TA, KII, 05]
>
> “…*Previously, we were providing the services under the tree. Now we have gotten tables, chairs, tents for shade and other tools from development partners including Amref Health Africa, but not enough to provide comprehensive services…” [DPP, WorHO, KII01]*

## Discussion

This study documented the integrated woreda-led MHS within the primary health care system, which has highlighted the contribution of a context-driven adaptive healthcare service delivery model for pastoral communities. The iterative participatory study design, process, and community-centred implementation of the MHS significantly improved access to essential maternal and child health services to communities that live in hard-to-reach areas. The MHS served as an entry mechanism to RMNCH-N health care services and strengthened awareness creation and local health facilities’ capacity development.

The MHS is an appropriate and imperative approach to improve access to and use of essential health services by people who live in remote areas in pastoral settings (15,31,32), which played a pivotal role in the move towards universal health coverage (21,31). The MHS improves access to services and uses convenient places in local communities in hard-to-reach areas (10,18). Furthermore, the MHS serves people who are displaced from their permanent residences due to natural and man-made disasters (33–35). The integrated MHS platform is an effective service delivery model designed using context-based participatory service arrangements to pastoralist communities who are vulnerable to seasonal and irregular shocks due to conflict and political insecurities (8,31,36). Similarly, adaptive alternative strategies have to be designed to deliver essential health services to pastoralist communities in the eastern African region, who have been facing social, environmental, economic, and political vulnerabilities (37). The MHS addresses gaps in low service utilisation of antenatal care, postnatal, immunisation, and family planning uptake in hard-to-reach rural areas of Ethiopia (38).

This MHS delivery model, iterative co-design and application of anchored community engagement enhanced the effectiveness of service implementation. It has demonstrated reaching marginalised people, pregnant women, under-five children, and elders who would not visit static health services for essential preventive, curative, and rehabilitative services. This finding was consistent with previous studies on mobile health services, which benefited remote settlements and hard-to-reach communities that have had no static health services. This remarkable achievement could be attributed to the commitment of Woreda health offices and the technical support of development partners. The MHS requires resources for progressive improvement to address the increasing demand of the community associated with demographic and epidemiological transitions (39,40). This study has highlighted that the implementation of MHS in pastoral settings strengthened local capacity, fostered community ownership, and built a resilient primary health care system (34). The community-centred MHS is crucial to establishing active emergency risk mitigation and early management of outbreaks associated with floods, conflict, and droughts (41).

This program learning has proved the importance of MHS in the pastoral context of Ethiopia, and the impact of active community engagement using VHLs who mobilise MHS beneficiaries from their villages to the point of care (42). The integration of nutritional intervention is a plus for the MHS to enhance acceptability by the residents (43,44). The MHS is a critical approach to ensure access to quality health services for women, girls, elders, and persons with disability where culture, beliefs, and values influence the use of essential health services in pastoralist communities (45,46). The MHS identified in this study demonstrates Woreda-led leadership commitment, PHCU-based participatory design, and effective community engagement and context-tailored communication, which have enhanced the ownership of MHS and fostered local capacity and ownership, a key strength of the study. The study used a multimethod approach for data acquisition; however, the study relied on two IPHCSD-supported woredas, service records, and its interpretation may need to consider its context.

## Implications

Health for all by 2000. It agreed upon a healthcare system which is universally accessible to individuals and acceptable to them, through their full participation, at a cost that the community and country can afford (47). Although it has been 25 years since the expected target year (2000), health remains inaccessible to all (47). Integrated MHS used community-centred, multidisciplinary, and active stakeholder involvement, using a tailored woreda-led approach (48), to foster institutionalisation and sustainability of MHS within the PHC system in the pastoral setting of Ethiopia. The MHS delivery model has a substantial contribution in ensuring access to essential health services (29) in the primary health care service delivery setting, where conventional health facilities are inaccessible to the community. It serves as one of the primary health care service delivery models to reach women, girls, elders, and persons with disability who are seriously affected by social determinants and abstain even from exempted services. The MHS delivery model is a mechanistic approach that needs substantial investment to contribute to and attain Sustainable Development Goal 3 and address the health, human rights, and development nexus. It is a novel approach to take health services to the community that reduces pressure on the facility-based healthcare delivery system sustainably in hard-to-reach areas.

## Conclusions

The integrated MHS delivery model is an innovative alternative service delivery strategy to ensure access to and use of essential health services in the pastoralist community in Ethiopia. It has shown substantial improvement in maternal and child health services, including contraceptive use, antenatal care coverage, immunisation, nutritional screening, and management of sick children. Women and girls made up the majority of the beneficiaries who are the most underserved through the existing routine health services. Although the woreda-led approach enhances the integration and suitability of the MHS within the primary health care system and fosters sustainability, the iMHS approach has also required innovative financing, supply refilling, and institutionalisation of financing mechanisms in the community-based health insurance scheme. The program should be accommodated within the existing health service delivery cost recovery mechanisms. It is recommended to scale up the Woreda-led integrated MHS delivery model to communities in remote and hard-to-reach areas, particularly in Ethiopia and other similar settings, to ensure access to essential health services.

## Acronyms

ANC: Antenatal Care
CAR: Contraceptive Acceptance Rate
HEW: Health Extension Workers
HEP: Health Extension Program
IPHCSD: Improve Primary health Care Services Delivery
KII: Key Informant Interview
LMIC: Low and Middle Income Countries
MCV: Measles Containing Vaccine
MHS: Mobile Health Service
MHNT: Mobile Health and Nutrition Team
PHC: Primary Health care
PHCU: Primary Health Care Unit
PNC: Post Natal Care
RMNHC: Reproductive Maternal Newborn Child Newborn Health
VHL: Village Health Leaders

## Declaration

### Consent for publication

Not applicable

### Availability of data and materials

All related data are presented fully within the paper and are available upon reasonable request to the lead author and the corresponding author.

## Acknowledgments

We would like to acknowledge the Gates Foundation for financial support, Amref Health Africa, and JSI for the overall administrative support, the study participants, data collectors, and supervisors for their willingness to give their time and information for this study.

## Supporting Information

S1 Datasets.

S2 Survey questionnaire

## Authors information

^1^Amref Health Africa in Ethiopia, Addis Ababa Ethiopia; ^2^JSI Research & Training Institute, Inc, Addis Ababa, Ethiopia; ^3^School of Public Health, Institute of Health Sciences, Jimma University, Jimma, Ethiopia; ^4^Ministry of Health, Addis Ababa, Ethiopia; and ^5^Gates Foundation, Ethiopia Country Office, Addis Ababa, Ethiopia; ^6^MERQ Consultancy P.L.C, Addis Ababa, Ethiopia

